# Post-stroke Innate Immune Dysfunction in Childhood Arterial Ischemic Stroke: Transcriptomic Signatures Distinguish Etiologies and Outcomes

**DOI:** 10.64898/2026.05.28.26354229

**Authors:** Mary C. Karalius, Prashanth S. Ramachandran, Maham Zia, Annie Wapniarski, Ravi Dandekar, Shiyin Wang, Nancy K. Hills, Huichun Xu, Max Wintermark, Nomazulu Dlamini, Marcela Torres, J. Michael Taylor, Sergio Baranzini, Joseph L. DeRisi, Heather J. Fullerton, Michael R. Wilson, VIPS II Investigators

## Abstract

**Background:** Immune-mediated mechanisms are increasingly implicated in childhood arterial ischemic stroke (AIS), but the associated inflammatory pathways and how they differ by stroke subtype and outcome remain poorly understood. Understanding immune responses to AIS may identify subtype-specific mechanisms and inform targeted strategies to reduce ischemic injury.

**Methods:** We conducted a prospective cohort study with cross-sectional transcriptomic analysis through the Vascular Effects of Infection in Pediatric Stroke Study Part II (VIPS II) at 22 academic centers in the United States, Canada, and Australia between December 2016 and January 2022. Children aged 28 days to 18 years with centrally confirmed AIS were enrolled within 72 hours of stroke onset, in addition to enrollment of stroke-free well children. Peripheral blood RNA sequencing was performed on samples collected within 72 hours of stroke or at enrollment for controls. Differential gene expression (DGE) and pathway analyses were performed comparing all AIS cases to stroke-free well children. Additional cross-sectional analyses stratified by stroke subtype and neurological outcomes were performed.

**Results:** Transcriptomes were available in 190/205 AIS cases (median age 11.7 years) and 91/100 stroke-free children (11.8 years). Stroke subtypes included 67 definite arteriopathic, 74 probable arteriopathic, 23 cardioembolic, and 26 idiopathic, with similar demographics but smaller infarct size for idiopathic cases. 47 genes (false discovery rate (FDR) <0.05 and log2 fold-change (log_2_FC)>1) were differentially expressed in AIS versus stroke-free well children, with upregulated pathways reflecting innate immune responses. Stratification by subtype revealed these inflammatory responses occurred after arteriopathic and cardioembolic AIS, but not idiopathic AIS; in sensitivity analyses, these findings were not explained by infarct size. Four immune-related genes were differentially expressed in children with good versus poor neurological outcomes at hospital discharge or 12 months; upregulation of one (Joining Chain; JCHAIN) correlated with poor outcomes at both timepoints.

**Conclusions:** Compared with stroke-free children, children with AIS, particularly arteriopathic and cardioembolic subtypes, have upregulated innate immune pathways, including neutrophil activation and interleukin-1 signaling. Differential expression of immune-related genes also correlated with neurological outcomes. These findings support immune dysregulation as a key feature of early pediatric AIS while highlighting differences across subtypes and clinical outcomes, with implications for targeted immunomodulatory therapies and future biomarker development.

## INTRODUCTION

Childhood arterial ischemic stroke (AIS) causes lifelong intellectual and physical disability with an annual global incidence of 1.3 per 100,000 children.^1^ Unlike adult AIS pathogenesis, driven by atherosclerosis and cardiovascular risk factors, childhood AIS is most commonly attributed to non-atherosclerotic arteriopathies, congenital heart disease, genetic disorders like sickle cell disease, and prothrombotic disease states; up to 25% of cases remain idiopathic.^2,3^ Our prior work found that routine childhood infections transiently increase risk of childhood AIS, while vaccinations are protective.^4–6^ These associations were seen across childhood stroke subtypes—arteriopathic, cardioembolic, idiopathic— suggesting that infection plays a role in different mechanistic pathways to AIS. The inflammatory response to infection could contribute to stroke pathogenesis through pro-thrombotic changes^7^ or injury to arterial or cardiac endothelium. Knowledge of the inflammatory pathways involved is needed for targeted approaches to both stroke prevention and post-stroke neuroprotection.

Differential gene expression (DGE) analyses of peripheral blood have shown promise in elucidating immune mechanisms in the immediate post-stroke period in adults, consistently demonstrating broad immune-related transcriptional changes following AIS.^8–15^ Additional DGE studies have distinguished between adult stroke subtypes, potentially reflecting distinct upstream inflammatory pathways, and some have further correlated transcriptomic signatures with clinical outcomes.^16–20^

To our knowledge, no prior study has investigated transcriptomic signatures in pediatric AIS. Here, we report VIPS II study results of peripheral gene expression profiling in a large, multinational cohort of children with AIS (within 72 hours of stroke ictus), compared to that in stroke-free well children. We tested the *a priori* hypotheses that children with AIS exhibit dysregulated inflammatory responses after their stroke, that their gene expression patterns differ by stroke subtype and that these profiles correlate with short- and long-term clinical outcomes.

## METHODS

This study is reported according to STROBE guidelines. All sites had ethics approval and obtained informed consent/assent for all enrollments.

### Setting and Patent Enrollment

We present a cross-sectional analysis of blood samples collected from the VIPS II prospective cohort of children (aged 28 days-18 years) with AIS, defined as an arterial infarct on brain imaging with corresponding acute neurological deficits. Setting, patient enrollment, and stroke subtype classification have been described elsewhere.^21^ Eligible patients were enrolled, and a blood sample was collected within 72 hours of stroke ictus (or last seen normal) at 22 sites in the U.S. (19), Canada (2), and Australia (1). To reduce heterogeneity, we excluded iatrogenic strokes and strokes that did not fall into the three most common subtypes: arteriopathic (definite or probable), cardioembolic, or idiopathic **(eMethods1)**. Sites collected clinical data via standardized chart abstraction forms (created by the International Pediatric Stroke Study (IPSS)) and patient/surrogate interviews (developed for the original VIPS study). VIPS II sites also enrolled and collected blood samples from stroke-free well children (defined as no clinical infection, acute hospitalization, or vaccinations in the prior two weeks) to serve as a reference for gene expression in well states independent of stroke **(Figure 1A)**.

**Figure 1.**
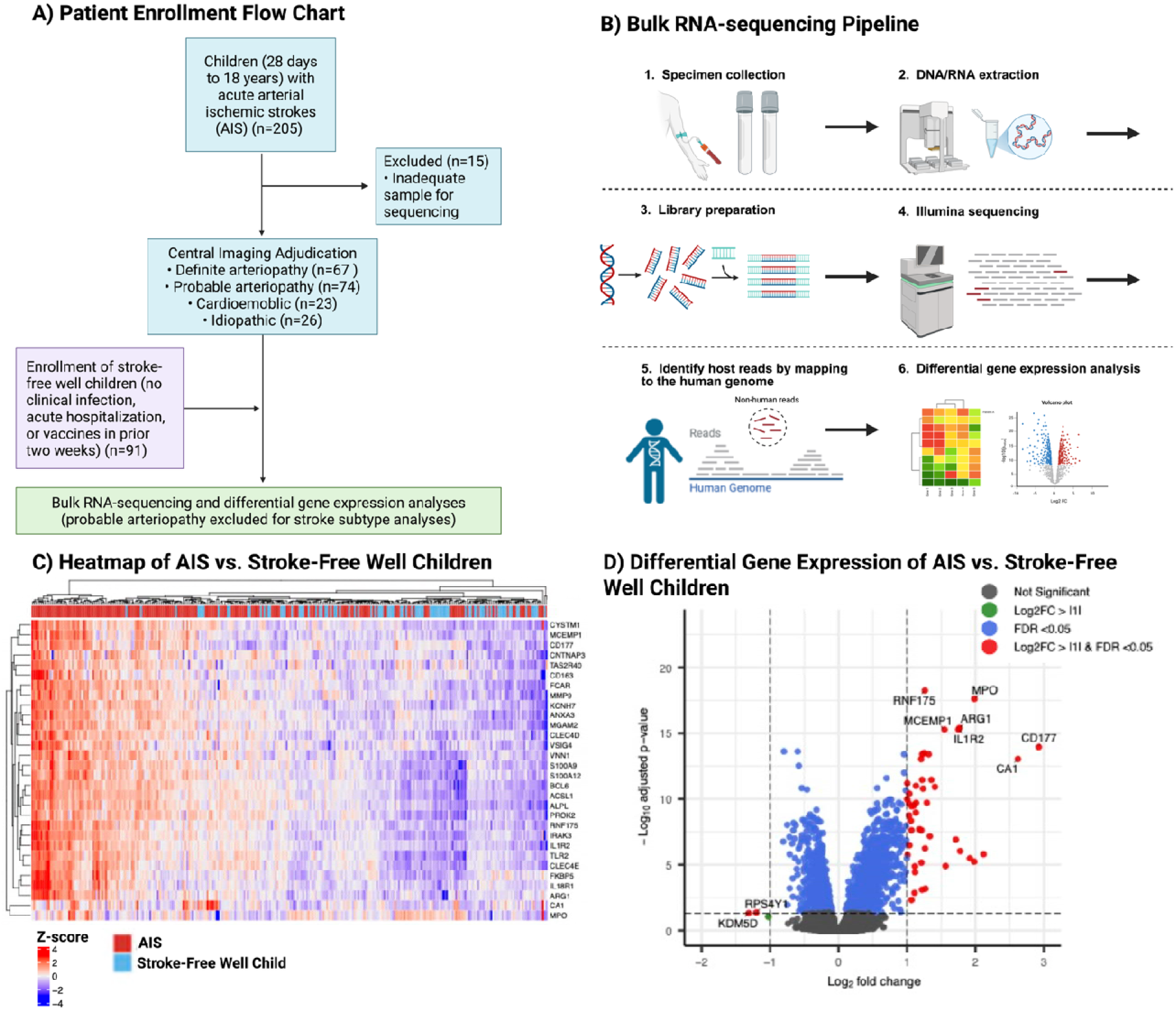
Methods and Differential Gene Expression Analysis between AIS and Stroke-Free Well Children. A) Flow chart of patient enrollment. B) Bulk RNA-sequencing and differential gene expression pipeline. C) Heatmap arranged by unsupervised clustering of top 30 differentially expressed genes between AIS patients and stroke-free well children. D) Volcano plot showing differentially expressed genes between AIS patients and stroke-free well children.

Neurological Outcomes: Site investigators classified neurological deficits at discharge as none, mild, moderate or severe. Twelve-month (±6 month) outcomes were assessed using the Pediatric Stroke Outcome Measure (PSOM) and dichotomized as “good” if the PSOM score was ≤1.5 and “poor” if the PSOM score was >1.5.^22^

## Study Specimens and RNA-Sequencing

Sites collected whole blood samples in PaxGene tubes (Ref No. 762165, Lot No. 1021321). Samples were centrifuged, and plasma was stored at −80°C at the site, then shipped to the University of California, San Francisco for central processing. RNA sequencing was performed via standard protocols as previously described **(Figure 1B).**^21^

### Data Analysis

#### Clinical Data

Clinical data for each patient were summarized using medians and interquartile ranges (IQR) for continuous variables and frequencies and percentages for categorical variables. Differences in demographics across the subtypes were compared using the Wilcoxon rank-sum test for continuous variables and the χ2 test (or Fisher exact, where appropriate) for categorical variables. Analyses were conducted using Stata v17.0 (Stata Corp, College Station, TX) with a 2-sided alpha set at 0.05.

#### Differential Gene Expression

We performed DGE analyses in all children with AIS compared to stroke-free children **(eMethods2)**. We then performed additional cross-sectional analyses stratified by stroke subtype, using stroke-free well children as the reference group for each analysis as well as comparison of stroke subtypes to each other. We then determined whether gene expression correlated with dichotomized neurological outcomes: none/mild versus moderate/severe deficits at discharge and good versus poor 12-month outcomes.

#### Pathway Analysis

Gene ontology (GO) and Gene Set Enrichment Analysis (GSEA) using clusterProfiler (version 4.14.6) in R were performed to investigate biologically meaningful pathways based on differentially expressed genes with liberalized logFC > or < 0 to improve sensitivity for pathway detection of more subtle changes.^27^ Pathway analysis was also performed using the Qiagen Ingenuity Pathway Analysis (IPA) toolkit.^28^ All pathways that were significant at BH p-adjusted <0.05 were considered.

## RESULTS

### Patient Cohort

The full patient cohort (N=205) has previously been described.^21^ Bulk RNA sequencing data were available for 190 cases (median age 11.7, interquartile range [IQR] 5.2-15.6 years; 45% female) and 91 stroke-free well children (11.8, 6.7-15.3 years; 53% female) (**Table 1**). Stroke subtypes of the 190 cases were definite arteriopathic (n=67), probable arteriopathic (n=74), cardioembolic (n=23), and idiopathic (n=26). Demographics and infectious exposure were similar between subtypes. Idiopathic cases had smaller infarct volumes than other subtypes and trended towards better short- and long-term outcomes (**Table 2**).

**Table 1:** Characteristics of childhood arterial ischemic stroke (AIS) cases and stroke-free children with RNA sequencing data.

| Characteristic | AIS Cases<br>(N=190) |  | Stroke-free, well<br>(N=91) |  |
| --- | --- | --- | --- | --- |
|  | n | (%) | n | (%) |
| **Demographics** |  |  |  |  |
| Age in years, median (IQR) | 11.7 | (5.2, 15.6) | 11.8 | (6.7, 15.3) |
| Sex, female | 85 | (44.7) | 48 | (52.7) |
| Race/ethnicity |  |  |  |  |
| White, non-Hispanic | 102 | (53.7) | 46 | (50.5) |
| Black, non-Hispanic | 23 | (12.1) | 10 | (11.0) |
| Asian | 15 | (7.9) | 4 | (4.4) |
| Hispanic | 41 | (21.6) | 14 | (15.4) |
| Native American/First Nations | 1 | (0.5) |  |  |
| Mixed or other | 7 | (3.7) | 12 | (13.2) |
| Unknown | 1 | (0.5) | 5 | (5.5) |
| **Socioeconomic Status** |  |  |  |  |
| Residence |  |  |  |  |
| Urban | 37 | (19.5) | 23 | (25.3) |
| Suburban | 111 | (58.4) | 58 | (63.7) |
| Rural | 41 | (21.6) | 16 | (17.6) |
| Missing | 1 | (0.5) |  |  |
| Household income (in US dollars) |  |  |  |  |
| <\$10,000 | 12 | (6.3) | 3 | (3.3) |
| \$10,000-30,000 | 24 | (12.6) | 8 | (8.8) |
| \$31,000-50,000 | 23 | (12.1) | 11 | (12.1) |
| \$50,000-100,000 | 43 | (22.6) | 21 | (23.1) |
| >100,000 | 54 | (28.4) | 46 | (50.5) |
| Missing | 34 | (17.9) | 2 | (2.2) |
| Maternal education, highest level |  |  |  |  |
| Less than high school | 13 | (6.8) | 1 | (1.1) |
| High school graduate | 44 | (23.2) | 14 | (15.4) |
| Some college education | 57 | (30.0) | 17 | (18.7) |
| Bachelor's degree, or equivalent | 38 | (20.0) | 24 | (26.4) |
| Some graduate education | 6 | (3.2) | 1 | (1.1) |
| Graduate degree | 29 | (15.3) | 34 | (37.4) |
| Missing | 3 | (1.6) | 34 | (37.4) |

**Table 2:**
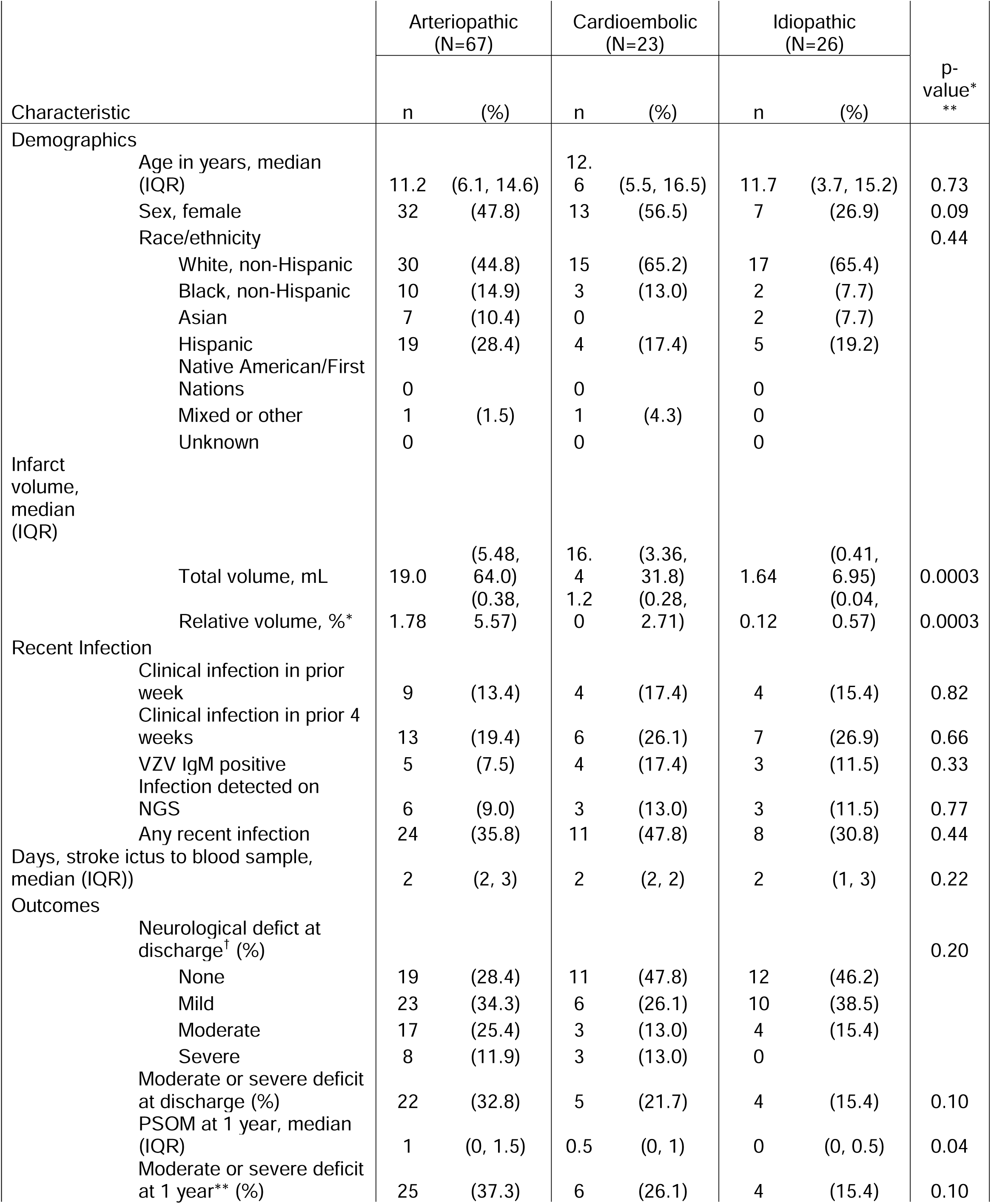
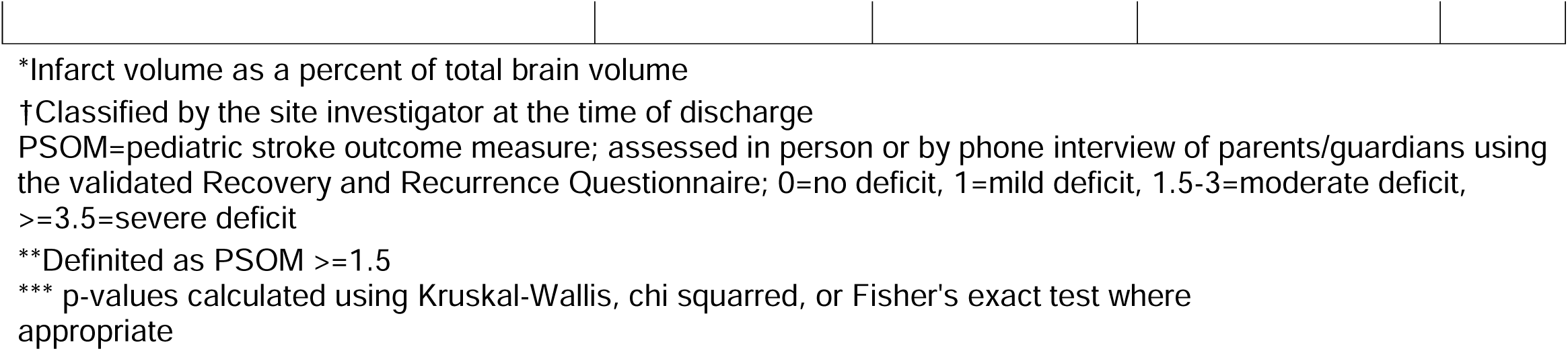
Characteristics of childhood arterial ischemic stroke (AIS) cases with RNA sequencing data, stratified by stroke type.

|  |  | Arteriopathic<br>(N=67) |  | Cardioembolic<br>(N=23) |  | Idiopathic<br>(N=26) |  | p-<br>value*<br>** |
| --- | --- | --- | --- | --- | --- | --- | --- | --- |
| Characteristic |  | n | (%) | n | (%) | n | (%) |  |
| Demographics |  |  |  |  |  |  |  |  |
|  | Age in years, median (IQR) | 11.2 | (6.1, 14.6) | 12.6 | (5.5, 16.5) | 11.7 | (3.7, 15.2) | 0.73 |
|  | Sex, female | 32 | (47.8) | 13 | (56.5) | 7 | (26.9) | 0.09 |
|  | Race/ethnicity |  |  |  |  |  |  | 0.44 |
|  | White, non-Hispanic | 30 | (44.8) | 15 | (65.2) | 17 | (65.4) |  |
|  | Black, non-Hispanic | 10 | (14.9) | 3 | (13.0) | 2 | (7.7) |  |
|  | Asian | 7 | (10.4) | 0 |  | 2 | (7.7) |  |
|  | Hispanic | 19 | (28.4) | 4 | (17.4) | 5 | (19.2) |  |
|  | Native American/First Nations | 0 |  | 0 |  | 0 |  |  |
|  | Mixed or other | 1 | (1.5) | 1 | (4.3) | 0 |  |  |
|  | Unknown | 0 |  | 0 |  | 0 |  |  |
| Infarct volume, median (IQR) |  |  |  |  |  |  |  |  |
|  | Total volume, mL | 19.0 | (5.48, 64.0) | 16.4 | (3.36, 31.8) | 1.64 | (0.41, 6.95) | 0.0003 |
|  | Relative volume, %* | 1.78 | (0.38, 5.57) | 1.2 | (0.28, 2.71) | 0.12 | (0.04, 0.57) | 0.0003 |
| Recent Infection |  |  |  |  |  |  |  |  |
|  | Clinical infection in prior week | 9 | (13.4) | 4 | (17.4) | 4 | (15.4) | 0.82 |
|  | Clinical infection in prior 4 weeks | 13 | (19.4) | 6 | (26.1) | 7 | (26.9) | 0.66 |
|  | VZV IgM positive | 5 | (7.5) | 4 | (17.4) | 3 | (11.5) | 0.33 |
|  | Infection detected on NGS | 6 | (9.0) | 3 | (13.0) | 3 | (11.5) | 0.77 |
|  | Any recent infection | 24 | (35.8) | 11 | (47.8) | 8 | (30.8) | 0.44 |
| Days, stroke ictus to blood sample, median (IQR)) |  | 2 | (2, 3) | 2 | (2, 2) | 2 | (1, 3) | 0.22 |
| Outcomes |  |  |  |  |  |  |  |  |
|  | Neurological deficit at discharge<sup>†</sup> (%) |  |  |  |  |  |  | 0.20 |
|  | None | 19 | (28.4) | 11 | (47.8) | 12 | (46.2) |  |
|  | Mild | 23 | (34.3) | 6 | (26.1) | 10 | (38.5) |  |
|  | Moderate | 17 | (25.4) | 3 | (13.0) | 4 | (15.4) |  |
|  | Severe | 8 | (11.9) | 3 | (13.0) | 0 |  |  |
|  | Moderate or severe deficit at discharge (%) | 22 | (32.8) | 5 | (21.7) | 4 | (15.4) | 0.10 |
|  | PSOM at 1 year, median (IQR) | 1 | (0, 1.5) | 0.5 | (0, 1) | 0 | (0, 0.5) | 0.04 |
|  | Moderate or severe deficit at 1 year** (%) | 25 | (37.3) | 6 | (26.1) | 4 | (15.4) | 0.10 |

### RNA-Sequencing and Data Pre-Processing

Median sequencing depth for the plasma RNA-seq libraries was 55,158,536 paired-end reads (IQR, 50,217,786 – 62,215,054). Transcripts mapped to a total of 60,591 genes and of those, transcripts aligning to the 19,263 protein-coding genes were used for downstream analyses. The median number of non-zero transcript protein-coding genes per sample was 17,768 (IQR 16,800-18,590). One extreme outlier with 10-60X fold higher gene transcripts compared to other cases was removed from the analyses.

### DGE and Pathway Analysis in AIS versus Stroke-free Well Children

#### DGE Analysis

Unsupervised hierarchical clustering demonstrated separation between AIS patients and stroke-free well children (**Figure 1C**). A total of 2,754 genes were differentially expressed (FDR <0.05), with 1,561 genes upregulated (log_2_FC>0) and 1,193 genes downregulated (log_2_FC<0). 47 genes had a log_2_FC > 1, and 2 genes had log_2_FC < -1 **(Figure 1D, eTable1**). Of the top 20 differentially expressed genes based on log_2_FC, 14 were related to innate immune function, including MPO, IL-1 receptor type 2 (IL-1R2), and CD177 (**eTable2**).

#### Pathway Analyses

GO pathway analysis of all significantly upregulated and downregulated genes identified 664 upregulated pathways and 82 downregulated pathways in AIS patients (**eFigure1-2, eTable3-4**). To complement this, GSEA, which considers all genes ranked by expression change, identified 348 differentially regulated pathways **(eFigure3, eTable5**). GSEA also highlighted upregulation of pathways involved in innate immunity, including neutrophil activation and interleukin-1 (IL-1)/Interleukin-8 (IL-8) signaling and downregulation of pathways involved in T-cell differentiation. IPA further highlighted dysregulation of key immune pathways, with the most significant being neutrophil degranulation **(Figure 2A).** IL-1 family signaling and toll-like receptor (TLR) signaling were also significant (**Figure 2A**). Network analysis demonstrated the complex interplay between these pathways with IL1B serving as a hub molecule (**Figure 2B**).

**Figure 2.**
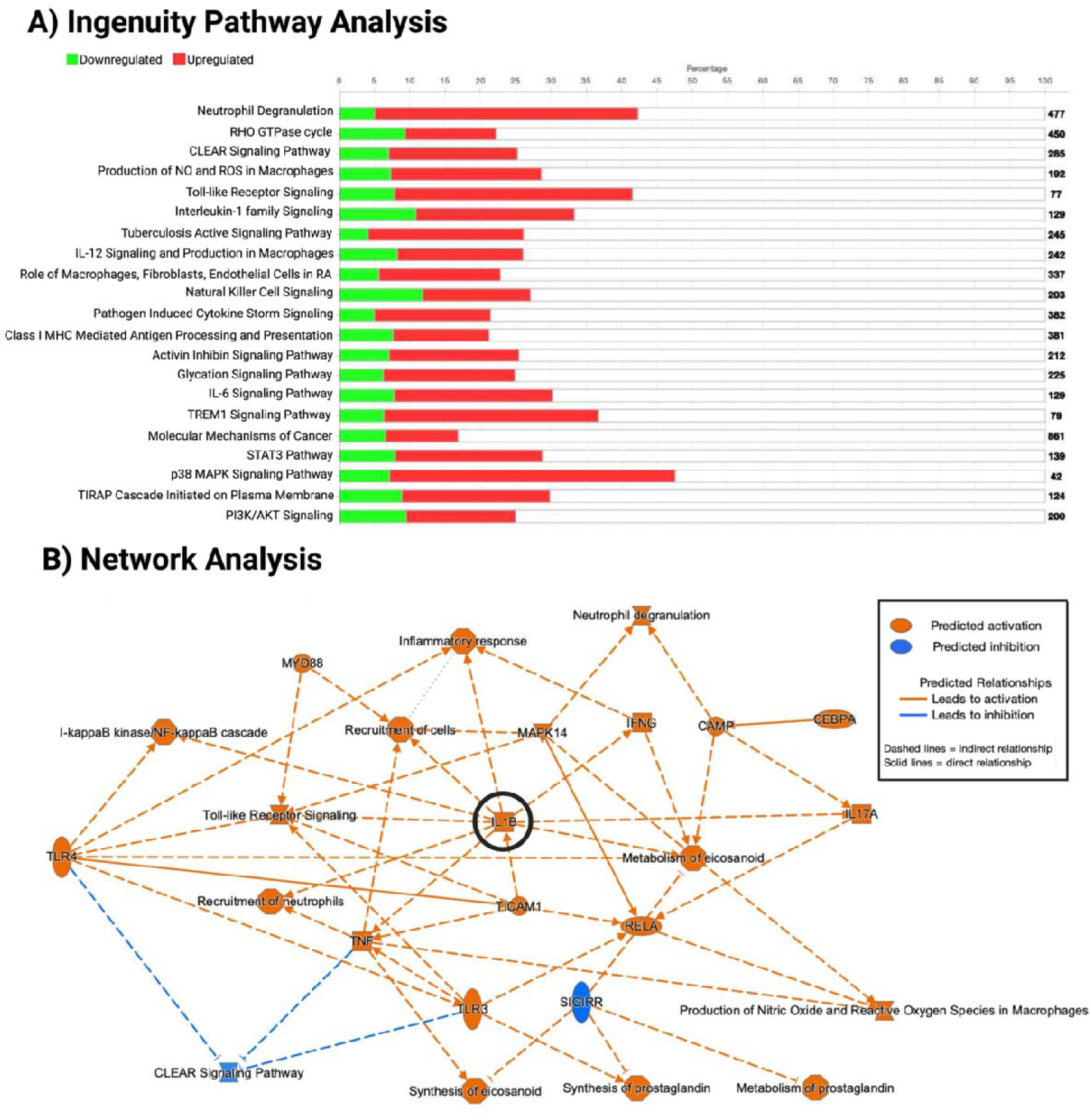
Pathway and Network Analyses of AIS versus Stroke-Free Well Children. A) Ingenuity Pathway Analysis (IPA) results demonstrating top differentially expressed pathways in AIS cases compared to stroke-free well children. The number on the far right is the total number of genes in the IPA pathway database. The red bar represents the percentage of significant upregulated genes, and the green bar represents the percentage of downregulated genes. **B)** Network analysis of significant upregulated pathways and genes demonstrating IL-1B as key hub molecular, with close relationship to activation of neutrophils and toll-like receptors.

### DGE and Pathway Analysis Stratified by Stroke Subtypes

#### Stroke Subtypes Compared to Stroke-Free Well Children

When comparing individual stroke subtypes to stroke-free well children, 95 genes were upregulated (and 1 gene downregulated) in arteriopathic cases, compared to 83 genes upregulated and 3 genes downregulated in cardioembolic cases, and only 1 gene upregulated and 0 genes downregulated in idiopathic stroke cases **(Figure 3A-D, eTable6-8)**. Only one gene was upregulated across all stroke subtypes versus stroke-free well children: CD177.

**Figure 3.**
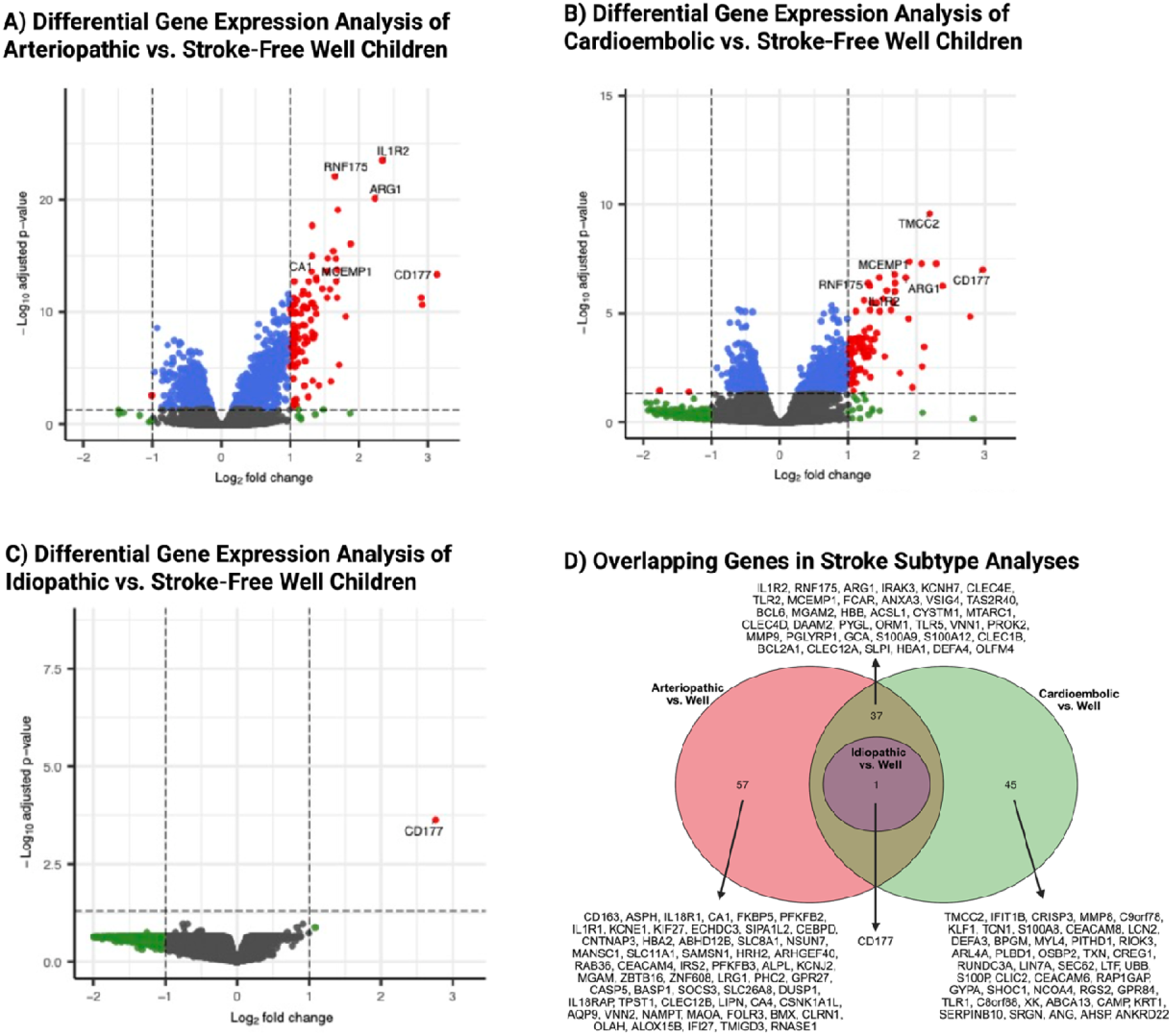
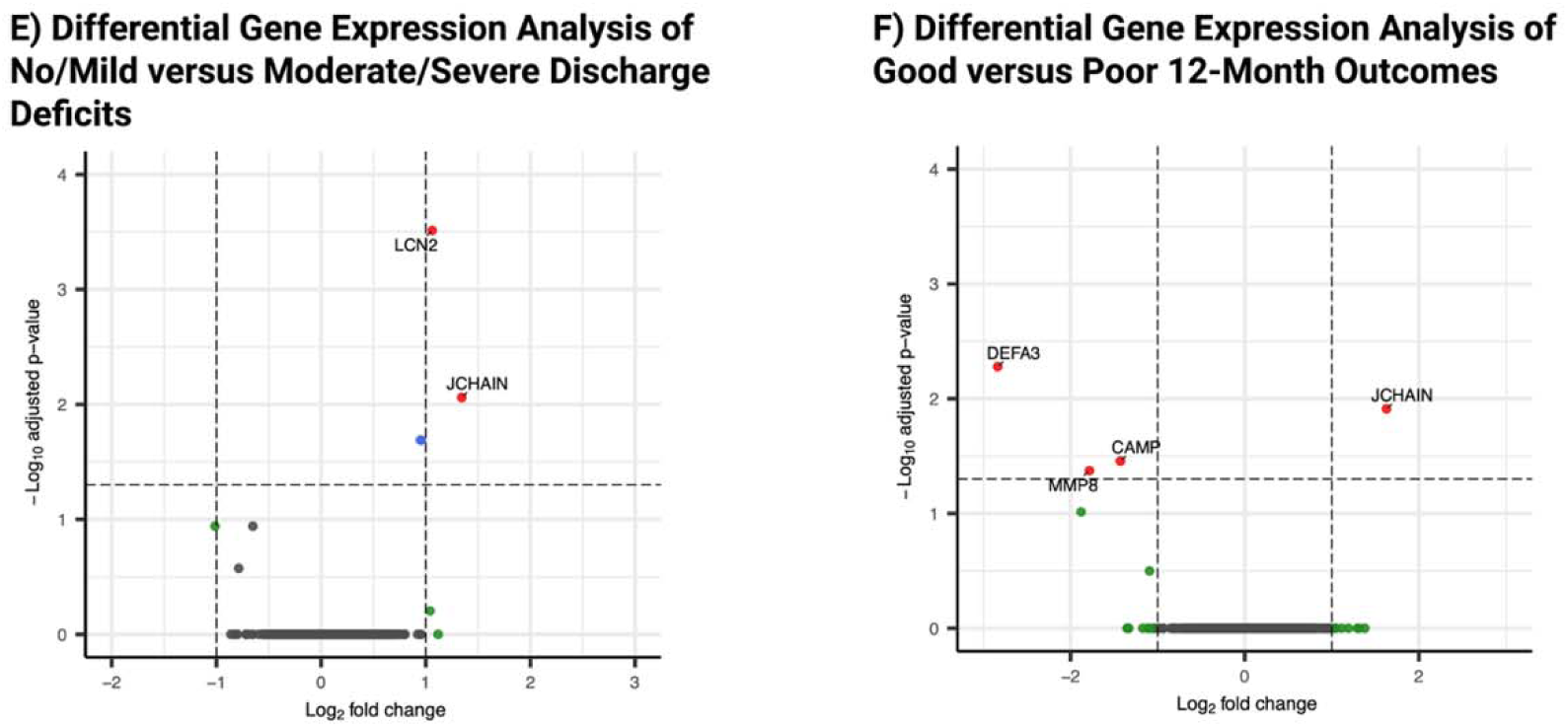
Differential Gene Expression Analysis by Stroke Subtype and Clinical Outcomes. Volcano plots of differentially expressed genes in children with AIS (vs. stroke-free well children) stratified by stroke subtype and outcome (A-C, E-F): A) Arteriopathic vs. Stroke-Free Well Children. B) Cardioembolic vs. Stroke-Free Well Children. C) Idiopathic vs. Stroke-Free Well Children. D) Venn Diagram of comparison of top differentially expressed genes when comparing stroke subtypes versus stroke-free well children. E) No/mild deficits vs. moderate/severe deficits at discharge. F) Good vs poor outcomes based on PSOM as 12-months.

GO pathway analyses of upregulated genes in arteriopathic cases (versus well children) and cardioembolic cases (versus well children) demonstrated upregulation of the innate immune system and IL-1, consistent with the overall AIS DGE analysis **(eFigure4-5, eTable9-10).**

#### Stroke Subtypes Compared to Each Other

When directly comparing gene expression between the stroke subtypes, we observed inflammatory gene upregulation in both the arteriopathic and cardioembolic cases, compared to idiopathic cases **(eTable11-12, eFigure 6-7)** whereas gene expression in arteriopathic versus cardioembolic cases was similar **(eTable13, eFigure8).**

### Sensitivity Analyses Related to Infarct Volume

We performed sensitivity analyses to control for the greater infarct volume in cases of arteriopathic versus idiopathic stroke. We compared the ten smallest arteriopathic strokes (relative median infarct volume [IQR] 0.06 [0.03-0.1]) to stroke-free well children and found a similar inflammatory gene expression pattern to our larger analysis **(eTable14).** A direct comparison of small-volume arteriopathic strokes to idiopathic strokes also demonstrated a similar inflammatory differential gene expression pattern **(eTable15).** Spearman’s rank correlations between relative infarct volume and individual gene expression levels also did not find that gene expression was correlated with infarct volume **(eTable16)**.

### DGE and Neurological Outcomes

Data on neurological deficits at discharge were available for all AIS cases, and 12-month PSOM scores were available for 136/189 cases (72%). Compared to children with no/mild deficits at discharge (n=120), those with moderate/severe deficits (n=69) demonstrated upregulation of two genes: lipocalin-2 (LCN2) and Joining CHAIN (JCHAIN) **(Figure 3E).** When comparing gene expression of AIS cases with good (n=97) versus poor (n=39) outcomes at 12-month follow-up, JCHAIN upregulation again correlated with poor outcomes, whereas upregulation of defensin alpha 3 (DEFA3), cathelicidin antimicrobial peptide (CAMP), and matrix metallopeptidases (MMP8) correlated with good outcomes **(Figure 3F).**

## DISCUSSION

To our knowledge, we present the first analysis of peripheral gene expression in children with AIS. We observed upregulation of numerous inflammatory genes following childhood AIS relative to stroke-free well children. Enriched biological pathways were most consistent with innate immune responses, particularly neutrophil activation and IL-1 signaling. Stratification by stroke subtype revealed that this response was driven by children with arteriopathic and cardioembolic stroke, but not idiopathic stroke. In addition, expression of two genes correlated with greater deficits at discharge, one of which also correlated with poorer 12-month outcomes (i.e., JCHAIN). In contrast, downregulation of three neutrophil-mediated genes was associated with favorable long-term outcomes.

Our top differentially expressed genes in pediatric AIS showed substantial overlap with prior adult AIS peripheral DGE studies.^12,14,29^ Barr et al. identified a 9-gene expression signature in adult AIS patients within 24 hours of symptom onset, of which MMP9, ARG1, S100A12 were also significantly upregulated in our cohort.^14^ These same genes were identified by Tang et al. who identified an 18-gene expression panel distinguishing acute adult AIS from non-stroke controls.^12^ Two additional genes from that panel, S100A9 and BCL6, were also significantly upregulated in our cohort.^12^ In a larger adult AIS cohort, Raman et al. similarly replicated 78% of genes identified by Barr et al. and 81% identified by Tang et al. Of their top 10 genes, three – MCEMP1, ANXA3, and IRAK3 – were among the topmost differentially expressed in our cohort. In addition, MMP8, which was upregulated in our cohort, was recently reported to be increased in children with tuberculous meningitis-associated stroke compared to those without stroke.^30^ Despite differences in patient age, sample timing, analytic methods, and clinical interventions, the consistent identification of several key immune-related genes across cohorts suggests a conserved inflammatory host response to AIS, supporting further investigation of these genes as candidate biomarkers and potential therapeutic targets.

CD177 expression, conserved across all subtype analyses, encodes a glycosyl-phosphatidylinositol (GPI)-linked cell surface protein, primarily expressed on neutrophils and is involved in neutrophil activation and migration. In an adult study, patients had increased peripheral CD177+ neutrophils following stroke compared to controls, while experimental mouse models of middle cerebral occlusion (MCAO) show increased infiltration of CD177+ neutrophils in areas of ischemic injury.^48^ Neutrophils are key mediators of early ischemic injury, thought to contribute to blood-brain barrier disruption and secondary tissue damage through release of proinflammatory cytokines and MMPs, followed by infiltration into ischemic tissue.^11^ Consistent with this, adult AIS cohorts have associated higher neutrophil counts with worse functional outcomes and increased mortality, while experimental models have demonstrated that targeting neutrophils may improve stroke outcomes.^11,32,33^ Furthermore, an experimental model of virally induced childhood arteriopathy implicated neutrophil-driven injury through activation of the TLR3-neutrophil axis, driving blood-brain barrier disruption.^34^ In our cohort, pathway analysis also demonstrated enrichment of TLR signaling pathways, particularly TLR3 and TLR4, which are important upstream regulators of innate immune activation and neutrophil responses after ischemic injury.^35,36^ Similar upregulation of TLR signaling has been reported in adult AIS studies.^14,15,20^ Collectively, these results support a central role for innate immune and neutrophil-mediated inflammatory responses in childhood AIS. Indeed, there are several early clinical trials targeting various aspects of neutrophil-mediated inflammation to promote stroke recovery.^11^

The IL-1 signaling pathway was also upregulated in our childhood AIS cases, and pathway analysis identified IL1B as key hub molecule, consistent with adult stroke studies.^15,20,37^ IL-1 is a key mediator of the innate immune response and activates neutrophils through the IL-1 receptor.^38^ IL1R2 was among our top 10 differentially expressed genes, with the highest expression observed in the arteriopathic subtype. IL-1R2 serves as a decoy receptor, exerting an anti-inflammatory response by blocking downstream IL-1 signaling pathways.^39^ We suspect that the increased expression of IL-1R2 may represent a compensatory response to limit excessive post-ischemic inflammation.^40,41^ Experimental mouse models have shown that treatment with recombinant IL-1 receptor antagonist (IL-1Ra) reduced ischemic injury following MCAO.^42,43^ Additionally, a phase 2 randomized controlled trial in adult AIS demonstrated that treatment with recombinant IL-1Ra was safe and well tolerated (primary aim) with secondary analyses suggesting improved clinical outcomes.^44^ Taken together, these findings support IL-1 blockade as another potential therapeutic strategy to mitigate early post-stroke inflammation.

Several macrophage-associated genes (CD163, ARG1, ALOX15B) were upregulated in arteriopathic but not cardioembolic stroke when compared directly to the idiopathic subtype. CD163 is a scavenger receptor involved in hemoglobin clearance after vascular injury and is also considered a canonical marker of alternatively activated (M2-like) macrophages which are involved in tissue repair and anti-inflammatory responses.^45,46^ Clinically, CD163+ macrophages are implicated in inflammatory vascular disorders including ANCA-associated renal vasculitis and giant cell arteritis.^47–50^ ALOX15B, a macrophage-expressed lipoxygenase that generates pro-inflammatory oxidized lipids, has also been implicated in arterial wall inflammation.^51^ Elevated ARG1, another marker of alternatively activated macrophages, has been linked to worse post-stroke functional recovery after adult AIS.^52,53^ Together, the coordinated upregulation of this set of genes suggests activation of macrophage-mediated vascular injury and/or repair mechanisms that are more pronounced in arteriopathic subtypes in comparison to cardioembolic AIS.

In contrast, children with idiopathic AIS demonstrated minimal inflammatory gene activation, and tended to have less disability at discharge, findings not explained by their smaller infarct volume. It is possible that inflammatory changes in this subgroup were either less robust or more transient than in arteriopathic or cardioembolic cases. The smallest arteriopathic cases demonstrated upregulation of inflammatory genes relative to stroke-free well children, further supporting the concept that factors beyond infarct size likely influence post-stroke immune activation and clinical outcomes.^54^

There was also notable downregulation of T-cell mediated pathways including T-cell selection and differentiation. While T-cell responses have been implicated in both protective and pathogenic processes after stroke, their precise role remains unclear.^55^ Prior studies suggest that certain T-cell populations, particularly CD4+ regulatory T cells (Tregs), may exert neuroprotective effects by limiting excessive inflammation and promoting tissue repair and long-term stroke recovery.^56–60^ In this context, the relative downregulation of T-cell associated pathways may reflect impaired adaptive immune regulation or a shift towards a predominantly innate inflammatory response in the acute post-stroke period.

We identified two genes that were upregulated in those who had worse deficits at hospital discharge: LCN2 and JCHAIN. LCN2 encodes an innate immune protein which has been increasingly implicated in ischemic stroke.^61^ Experimental models demonstrate rapid upregulation of LCN2 in the post-ischemic brain, where it may contribute to ongoing injury through promotion of neuroinflammation, glial activation, blood–brain barrier disruption, and neuronal cell death, while LCN2 deficiency may attenuate infarct severity and neurological impairment.^62,63^ In adults with AIS, elevated LCN2 has also been associated with greater stroke severity at stroke onset and worse functional 90-day outcomes.^63–65^ In contrast, DEFA3, CAMP, and MMP8, which are involved in neutrophil activation and degranulation, were upregulated in children with better long-term outcomes, raising the possibility that certain innate immune responses may instead reflect protective or reparative processes involved in recovery and tissue remodeling.

JCHAIN upregulation at baseline also correlated with poor outcomes at 12 months post-stroke. JCHAIN, which is involved in humoral immune responses through its role in IgA and IgM polymerization, has been less well studied in ischemic stroke.^66^ This finding may suggest a potential deleterious contribution of adaptive immune activation to pediatric AIS outcomes, but warrants further investigation.

## LIMITATIONS

Our study has several limitations. Our samples were collected within 72 hours post-AIS, and peripheral gene expression can change even over the course of hours. Similarly, given the cross-sectional study design and that samples were collected post-stroke, we cannot definitively establish whether differentially expressed genes reflect differences in stroke pathogenesis (e.g., inflammation prior to stroke onset) versus the host response to ischemia. Given the smaller sample size of the idiopathic stroke cohort and the likely increased heterogeneity of stroke etiologies in this group, there is also the possibility that we were underpowered to detect more subtle changes in DGE for idiopathic AIS. Our outcomes data were limited given that the IPSS data collection forms have no established criteria for the classification of deficits at discharge; this variable reflects investigator perceptions and may vary by investigator and site. In addition, we were missing 12-month PSOM data from a quarter of our cases, and the PSOM emphasizes motor deficits over deficits in other realms.^22^

## CONCLUSION

This is the first study to evaluate gene expression signatures in the immediate post-stroke period in a large cohort of children with AIS. We found evidence of innate immune dysregulation after pediatric AIS, with a complex interplay between several key proinflammatory pathways including neutrophils, IL-1 signaling, and TLR signaling. This inflammatory response was driven by the arteriopathic and cardioembolic stroke cases, as opposed to the idiopathic stroke, suggesting biological differences between stroke types with implications for future therapeutic targets for both stroke prevention and recovery. Furthermore, several genes were associated with clinical outcome, which may inform work on pediatric stroke recovery, such as serving as the basis for future biomarker development and prognostication.

## Data Availability

All data produced in the present study will be uploaded to GEO.

## ACKNOWLEDGEMENTS

The authors wish to acknowledge the important contributions of the research coordinators at the VIPS II study (Vascular Effects of Infection in Pediatric Stroke) sites, and the VIPS II project managers, Maria Kuchherzki and Kathleen Colao, the patients and their families.

## VIPS II INVESTIGATORS

Michael Dowling, Christine Fox, Nomazulu Dlamini, Gabrielle DeVeber, Marcela Torres, Jenny Wilson, Sarah Lee, Dana Cummings, Warren Lo, Melissa Chung, Lori Jordan, Tim Bernard, Megan Barry, Rebecca Ichord, Lauren Beslow, Mukta Sharma, Shannon Carpenter, Catherine Amlie-Lefond, Neil Friedman, John Michael Taylor, Michael Rivkin, Laura Lehman, Paola Pergami, Andrea Pardo, Tracee Ridley-Pryor, Ryan Felling, Lisa Sun, Mark Mackay, Adam Kirton.

## REFERENCES

1. Gao L, Lim M, Nguyen D, et al. The incidence of pediatric ischemic stroke: A systematic review and meta-analysis. Int J Stroke Off J Int Stroke Soc. 2023;18(7):765–772. doi:10.1177/17474930231155336

2. Wintermark M, Hills NK, DeVeber GA, et al. Clinical and Imaging Characteristics of Arteriopathy Subtypes in Children with Arterial Ischemic Stroke: Results of the VIPS Study. AJNR Am J Neuroradiol. 2017;38(11):2172–2179. doi:10.3174/ajnr.A5376

3. Wintermark M, Hills NK, DeVeber GA, et al. Arteriopathy Diagnosis in Childhood Arterial Ischemic Stroke Results of the VIPS Study. Stroke J Cereb Circ. 2014;45(12):3597–3605. doi:10.1161/STROKEAHA.114.007404

4. Fullerton HJ, Hills NK, Elkind MSV, et al. Infection, vaccination, and childhood arterial ischemic stroke: Results of the VIPS study. Neurology. 2015;85(17):1459–1466. doi:10.1212/WNL.0000000000002065

5. Hills NK, Sidney S, Fullerton HJ. Timing and number of minor infections as risk factors for childhood arterial ischemic stroke. Neurology. 2014;83(10):890–897. doi:10.1212/WNL.0000000000000752

6. Tudorache R, Jaboyedoff M, Gabet A, et al. Infection and Pediatric Arterial Ischemic Stroke Presumably Related to Focal Cerebral Arteriopathy: Data From the COVID-19 Pandemic. Stroke. 2024;55(6):1672–1675. doi:10.1161/STROKEAHA.123.045632

7. Elkind MSV, Boehme AK, Smith CJ, Meisel A, Buckwalter MS. Infection as a Stroke Risk Factor and Determinant of Outcome After Stroke. Stroke. 2020;51(10):3156–3168. doi:10.1161/STROKEAHA.120.030429

8. Stamova B, Knepp B, Rodriguez F. Molecular heterogeneity in human stroke - What can we learn from the peripheral blood transcriptome? J Cereb Blood Flow Metab Off J Int Soc Cereb Blood Flow Metab. Published online March 13, 2025:271678X251322598. doi:10.1177/0271678X251322598

9. Jayaraj RL, Azimullah S, Beiram R, Jalal FY, Rosenberg GA. Neuroinflammation: friend and foe for ischemic stroke. J Neuroinflammation. 2019;16(1):142. doi:10.1186/s12974-019-1516-2

10. Qin C, Yang S, Chu YH, et al. Signaling pathways involved in ischemic stroke: molecular mechanisms and therapeutic interventions. Signal Transduct Target Ther. 2022;7(1):215. doi:10.1038/s41392-022-01064-1

11. Jickling GC, Liu D, Ander BP, Stamova B, Zhan X, Sharp FR. Targeting neutrophils in ischemic stroke: translational insights from experimental studies. J Cereb Blood Flow Metab Off J Int Soc Cereb Blood Flow Metab. 2015;35(6):888–901. doi:10.1038/jcbfm.2015.45

12. Tang Y, Xu H, Du XL, et al. Gene Expression in Blood Changes Rapidly in Neutrophils and Monocytes after Ischemic Stroke in Humans: A Microarray Study. J Cereb Blood Flow Metab. 2006;26(8):1089–1102. doi:10.1038/sj.jcbfm.9600264

13. Grond-Ginsbach C, Hummel M, Wiest T, et al. Gene expression in human peripheral blood mononuclear cells upon acute ischemic stroke. J Neurol. 2008;255(5):723–731. doi:10.1007/s00415-008-0784-z

14. Barr TL, Conley Y, Ding J, et al. Genomic biomarkers and cellular pathways of ischemic stroke by RNA gene expression profiling. Neurology. 2010;75(11):1009–1014. doi:10.1212/WNL.0b013e3181f2b37f

15. Raman K, O’Donnell MJ, Czlonkowska A, et al. Peripheral Blood MCEMP1 Gene Expression as a Biomarker for Stroke Prognosis. Stroke. 2016;47(3):652–658. doi:10.1161/STROKEAHA.115.011854

16. Xu H, Tang Y, Liu DZ, et al. Gene expression in peripheral blood differs after cardioembolic compared with large-vessel atherosclerotic stroke: biomarkers for the etiology of ischemic stroke. J Cereb Blood Flow Metab Off J Int Soc Cereb Blood Flow Metab. 2008;28(7):1320–1328. doi:10.1038/jcbfm.2008.22

17. Jickling GC, Xu H, Stamova B, et al. Signatures of cardioembolic and large-vessel ischemic stroke. Ann Neurol. 2010;68(5):681–692. doi:10.1002/ana.22187

18. Jickling GC, Stamova B, Ander BP, et al. Profiles of lacunar and nonlacunar stroke. Ann Neurol. 2011;70(3):477–485. doi:10.1002/ana.22497

19. Jickling GC, Stamova B, Ander BP, et al. Prediction of cardioembolic, arterial, and lacunar causes of cryptogenic stroke by gene expression and infarct location. Stroke. 2012;43(8):2036–2041. doi:10.1161/STROKEAHA.111.648725

20. Amini H, Knepp B, Rodriguez F, et al. Early peripheral blood gene expression associated with good and poor 90-day ischemic stroke outcomes. J Neuroinflammation. 2023;20(1):13. doi:10.1186/s12974-022-02680-y

21. Karalius MC, Ramachandran PS, Wapniarski A, et al. Infection in Childhood Arterial Ischemic Stroke: Metagenomic Next-Generation Sequencing Results of the VIPS II Study. Stroke. 0(0). doi:10.1161/STROKEAHA.124.050548

22. Jordan LC, Hills NK, Fox CK, et al. Socioeconomic determinants of outcome after childhood arterial ischemic stroke. Neurology. 2018;91(6):e509–e516. doi:10.1212/WNL.0000000000005946

23. Deng Z, Delwart E. ContigExtender: a new approach to improving de novo sequence assembly for viral metagenomics data. BMC Bioinformatics. 2021;22(1):119. doi:10.1186/s12859-021-04038-2

24. Dobin A, Davis CA, Schlesinger F, et al. STAR: ultrafast universal RNA-seq aligner. Bioinforma Oxf Engl. 2013;29(1):15–21. doi:10.1093/bioinformatics/bts635

25. Ramachandran PS, Ramesh A, Creswell FV, et al. Integrating central nervous system metagenomics and host response for diagnosis of tuberculosis meningitis and its mimics. Nat Commun. 2022;13:1675. doi:10.1038/s41467-022-29353-x

26. Langelier C, Kalantar KL, Moazed F, et al. Integrating host response and unbiased microbe detection for lower respiratory tract infection diagnosis in critically ill adults. Proc Natl Acad Sci. 2018;115(52):E12353–E12362. doi:10.1073/pnas.1809700115

27. Yu G, Wang LG, Han Y, He QY. clusterProfiler: an R package for comparing biological themes among gene clusters. Omics J Integr Biol. 2012;16(5):284–287. doi:10.1089/omi.2011.0118

28. Krämer A, Green J, Pollard J, Tugendreich S. Causal analysis approaches in Ingenuity Pathway Analysis. Bioinforma Oxf Engl. 2014;30(4):523–530. doi:10.1093/bioinformatics/btt703

29. Peripheral Blood MCEMP1 Gene Expression as a Biomarker for Stroke Prognosis | Stroke. Accessed June 15, 2025. https://www-ahajournals-org.ucsf.idm.oclc.org/doi/10.1161/strokeaha.115.011854

30. Huynh J, Pretorius PM, Jan W, et al. Brain imaging and whole blood targeted transcriptomic analyses to characterise cerebral infarctions in children with tuberculous meningitis. Accessed August 24, 2025. 10.1093/infdis/jiaf399

31. Huang T, Xie W, Guo Y, et al. St3gal5-mediated sialylation of glyco-CD177 on neutrophils restricts neuroinflammation following CNS injury. Proc Natl Acad Sci. 2025;122(16):e2426187122. doi:10.1073/pnas.2426187122

32. Maestrini I, Tagzirt M, Gautier S, et al. Analysis of the association of MPO and MMP-9 with stroke severity and outcome. Neurology. 2020;95(1):e97–e108. doi:10.1212/WNL.0000000000009179

33. Buck BH, Liebeskind DS, Saver JL, et al. Early neutrophilia is associated with volume of ischemic tissue in acute stroke. Stroke. 2008;39(2):355–360. doi:10.1161/STROKEAHA.107.490128

34. Rayasam A, Jullienne A, Chumak T, et al. Viral mimetic triggers cerebral arteriopathy in juvenile brain via neutrophil elastase and NETosis. J Cereb Blood Flow Metab Off J Int Soc Cereb Blood Flow Metab. 2021;41(12):3171–3186. doi:10.1177/0271678X211032737

35. Tang SC, Arumugam TV, Xu X, et al. Pivotal role for neuronal Toll-like receptors in ischemic brain injury and functional deficits. Proc Natl Acad Sci. 2007;104(34):13798–13803. doi:10.1073/pnas.0702553104

36. Caso JR, Pradillo JM, Hurtado O, Lorenzo P, Moro MA, Lizasoain I. Toll-Like Receptor 4 Is Involved in Brain Damage and Inflammation After Experimental Stroke. Circulation. 2007;115(12):1599–1608. doi:10.1161/CIRCULATIONAHA.106.603431

37. Oh SH, Kim OJ, Shin DA, et al. Alteration of immunologic responses on peripheral blood in the acute phase of ischemic stroke: blood genomic profiling study. J Neuroimmunol. 2012;249(1-2):60–65. doi:10.1016/j.jneuroim.2012.04.005

38. Weber A, Wasiliew P, Kracht M. Interleukin-1 (IL-1) Pathway. Sci Signal. Published online January 19, 2010. doi:10.1126/scisignal.3105cm1

39. Peters VA, Joesting JJ, Freund GG. IL-1 receptor 2 (IL-1R2) and its role in immune regulation. Brain Behav Immun. 2013;32:1–8. doi:10.1016/j.bbi.2012.11.006

40. Li Q, Zhang X, Zhang Y, et al. Using proteomic biomarkers to estimate acute ischaemic stroke onset time. Commun Med. 2025;5(1):183. doi:10.1038/s43856-025-00895-7

41. Zhou Q, Dong Y, Wang K, Wang Z, Ma B, Yang B. A comprehensive analysis of the hub genes for oxidative stress in ischemic stroke. Front Neurosci. 2023;17. doi:10.3389/fnins.2023.1166010

42. Banwell V, Sena ES, Macleod MR. Systematic review and stratified meta-analysis of the efficacy of interleukin-1 receptor antagonist in animal models of stroke. J Stroke Cerebrovasc Dis Off J Natl Stroke Assoc. 2009;18(4):269–276. doi:10.1016/j.jstrokecerebrovasdis.2008.11.009

43. Sobowale OA, Parry-Jones AR, Smith CJ, Tyrrell PJ, Rothwell NJ, Allan SM. Interleukin-1 in Stroke. Stroke. 2016;47(8):2160–2167. doi:10.1161/STROKEAHA.115.010001

44. Emsley HCA, Smith CJ, Georgiou RF, et al. A randomised phase II study of interleukin-1 receptor antagonist in acute stroke patients. J Neurol Neurosurg Psychiatry. 2005;76(10):1366–1372. doi:10.1136/jnnp.2004.054882

45. Kristiansen M, Graversen JH, Jacobsen C, et al. Identification of the haemoglobin scavenger receptor. Nature. 2001;409(6817):198–201. doi:10.1038/35051594

46. Etzerodt A, Moestrup SK. CD163 and inflammation: biological, diagnostic, and therapeutic aspects. Antioxid Redox Signal. 2013;18(17):2352–2363. doi:10.1089/ars.2012.4834

47. Sakamoto A, Grogan A, Kawakami R, et al. Role of Hemoglobin-Stimulated Macrophages and Intraplaque Hemorrhage in the Development of Vascular Diseases. Arterioscler Thromb Vasc Biol. 2025;45(7):1021–1030. doi:10.1161/ATVBAHA.125.321439

48. Moran SM, Scott J, Clarkson MR, et al. The Clinical Application of Urine Soluble CD163 in ANCA-Associated Vasculitis. J Am Soc Nephrol JASN. 2021;32(11):2920–2932. doi:10.1681/ASN.2021030382

49. Estupiñán-Moreno E, Ortiz-Fernández L, Li T, et al. Methylome and transcriptome profiling of giant cell arteritis monocytes reveals novel pathways involved in disease pathogenesis and molecular response to glucocorticoids. Ann Rheum Dis. 2022;81(9):1290–1300. doi:10.1136/annrheumdis-2022-222156

50. O’Connell GC, Tennant CS, Lucke-Wold N, et al. Monocyte-lymphocyte cross-communication via soluble CD163 directly links innate immune system activation and adaptive immune system suppression following ischemic stroke. Sci Rep. 2017;7(1):12940. doi:10.1038/s41598-017-13291-6

51. Magnusson LU, Lundqvist A, Karlsson MN, et al. Arachidonate 15-Lipoxygenase Type B Knockdown Leads to Reduced Lipid Accumulation and Inflammation in Atherosclerosis. PLOS ONE. 2012;7(8):e43142. doi:10.1371/journal.pone.0043142

52. Supino D, Davoudian S, Silva-Gomes R, et al. Monocyte-macrophage membrane expression of IL-1R2 is a severity biomarker in sepsis. Cell Death Dis. 2025;16(1):269. doi:10.1038/s41419-025-07597-x

53. Kim HS, Jee SA, Einisadr A, et al. Detrimental influence of Arginase-1 in infiltrating macrophages on poststroke functional recovery and inflammatory milieu. Proc Natl Acad Sci. 2025;122(7):e2413484122. doi:10.1073/pnas.2413484122

54. Imaging Predictors of Neurologic Outcome After Pediatric Arterial Ischemic Stroke | Stroke. Accessed May 13, 2026. https://www-ahajournals-org.ucsf.idm.oclc.org/doi/10.1161/STROKEAHA.120.030965?url_ver=Z39.88-2003&rfr_id=ori:rid:crossref.org&rfr_dat=cr_pub%20%200pubmed

55. Functional role of regulatory lymphocytes in stroke: facts and controversies - PubMed. Accessed April 30, 2026. https://pubmed-ncbi-nlm-nih-gov.ucsf.idm.oclc.org/25791715/

56. Li N, Wang H, Hu C, Qie S, Liu Z. Regulatory T Cells for Stroke Recovery: A Promising Immune Therapeutic Strategy. CNS Neurosci Ther. 2025;31(1):e70248. doi:10.1111/cns.70248

57. Wang H, Wang Z, Wu Q, Yuan Y, Cao W, Zhang X. Regulatory T cells in ischemic stroke. CNS Neurosci Ther. 2021;27(6):643–651. doi:10.1111/cns.13611

58. Zhang Y, Liesz A, Li P. Coming to the Rescue: Regulatory T Cells for Promoting Recovery After Ischemic Stroke. Stroke. 2021;52(12):e837–e841. doi:10.1161/STROKEAHA.121.036072

59. Ito M, Komai K, Mise-Omata S, et al. Brain regulatory T cells suppress astrogliosis and potentiate neurological recovery. Nature. 2019;565(7738):246–250. doi:10.1038/s41586-018-0824-5

60. Shi L, Sun Z, Su W, et al. Treg cell-derived osteopontin promotes microglia-mediated white matter repair after ischemic stroke. Immunity. 2021;54(7):1527–1542.e8. doi:10.1016/j.immuni.2021.04.022

61. Zhao RY, Wei PJ, Sun X, et al. Role of lipocalin 2 in stroke. Neurobiol Dis. 2023;179:106044. doi:10.1016/j.nbd.2023.106044

62. Jin M, Kim JH, Jang E, et al. Lipocalin-2 deficiency attenuates neuroinflammation and brain injury after transient middle cerebral artery occlusion in mice. J Cereb Blood Flow Metab. 2014;34(8):1306–1314. doi:10.1038/jcbfm.2014.83

63. Hochmeister S, Engel O, Adzemovic MZ, et al. Lipocalin-2 as an Infection-Related Biomarker to Predict Clinical Outcome in Ischemic Stroke. PLOS ONE. 2016;11(5):e0154797. doi:10.1371/journal.pone.0154797

64. Keshk WA, Zineldeen DH, El-heneedy YA, Ghali AA. Thrombomodulin, alarmin signaling, and copeptin: cross-talk between obesity and acute ischemic stroke initiation and severity in Egyptians. Neurol Sci. 2018;39(6):1093–1104. doi:10.1007/s10072-018-3396-0

65. Huţanu A, Iancu M, Bălaşa R, Maier S, Dobreanu M. Predicting functional outcome of ischemic stroke patients in Romania based on plasma CRP, sTNFR-1, D-Dimers, NGAL and NSE measured using a biochip array. Acta Pharmacol Sin. 2018;39(7):1228–1236. doi:10.1038/aps.2018.26

66. Zhao J, Chen W, Li L, Zhang Z, Wang Y. JCHAIN: A Prognostic Marker Based on Pan-Cancer Analysis to Inhibit Breast Cancer Progression. Genes. 2025;16(9):1070. doi:10.3390/genes16091070

67. Kim HJ, Wei Y, Wojtkiewicz GR, Lee JY, Moskowitz MA, Chen JW. Reducing myeloperoxidase activity decreases inflammation and increases cellular protection in ischemic stroke. J Cereb Blood Flow Metab Off J Int Soc Cereb Blood Flow Metab. 2019;39(9):1864–1877. doi:10.1177/0271678X18771978

68. López-Bermejo A, Chico-Julià B, Castro A, et al. Alpha Defensins 1, 2, and 3. Arterioscler Thromb Vasc Biol. 2007;27(5):1166–1171. doi:10.1161/ATVBAHA.106.138594

69. Maneerat Y, Prasongsukarn K, Benjathummarak S, Dechkhajorn W, Chaisri U. Increased alpha-defensin expression is associated with risk of coronary heart disease: a feasible predictive inflammatory biomarker of coronary heart disease in hyperlipidemia patients. Lipids Health Dis. 2016;15(1):117. doi:10.1186/s12944-016-0285-5

70. Fernandez-Cadenas I, del Rio-Espinola A, Domingues-Montanari S, et al. Genes Involved in Hemorrhagic Transformations that Follow Recombinant T-Pa Treatment in Stroke Patients. Pharmacogenomics. 2013;14(5):495–504. doi:10.2217/pgs.13.19

71. Zhao M, Liu A, Wu J, Mo L, Lu F, Wan G. Il1r2 and Tnfrsf12a in transcranial magnetic stimulation effect of ischemic stroke via bioinformatics analysis. Medicine (Baltimore). 2024;103(4):e36109. doi:10.1097/MD.0000000000036109

72. Lakhan SE, Kirchgessner A, Tepper D, Leonard A. Matrix Metalloproteinases and Blood-Brain Barrier Disruption in Acute Ischemic Stroke. Front Neurol. 2013;4:32. doi:10.3389/fneur.2013.00032

73. Lindsberg PJ, Strbian D, Karjalainen-Lindsberg ML. Mast cells as early responders in the regulation of acute blood-brain barrier changes after cerebral ischemia and hemorrhage. J Cereb Blood Flow Metab Off J Int Soc Cereb Blood Flow Metab. 2010;30(4):689–702. doi:10.1038/jcbfm.2009.282

74. Wakisaka Y, Ago T, Kamouchi M, et al. Plasma S100A12 is associated with functional outcome after ischemic stroke: Research for Biomarkers in Ischemic Stroke. J Neurol Sci. 2014;340(1):75–79. doi:10.1016/j.jns.2014.02.031

75. Cheng MY, Lee AG, Culbertson C, et al. Prokineticin 2 is an endangering mediator of cerebral ischemic injury. Proc Natl Acad Sci U S A. 2012;109(14):5475–5480. doi:10.1073/pnas.1113363109

76. Liu L, Cai Y, Deng C. Identification of ANXA3 as a biomarker associated with pyroptosis in ischemic stroke. Eur J Med Res. 2023;28:596. doi:10.1186/s40001-023-01564-y

77. Zhang Z, Zhang M, Li D, et al. Microglial Annexin A3 Downregulation Alleviates Ischemic Injury by Inhibiting NF-κB/NLRP3-mediated Inflammation. Inflammation. Published online March 15, 2025:1–12. doi:10.1007/s10753-025-02287-4

