## Supplemental Methods and Figures for "Post-stroke Innate Immune Dysfunction in Childhood Arterial Ischemic Stroke: Transcriptomic Signatures Distinguish Etiologies and Outcomes"

**eMethods1: Classification of Stroke Subtypes**

A pediatric stroke neurologist (HJF) and neuroradiologist (MW) centrally reviewed all baseline and follow-up brain/vascular imaging and clinical data to confirm the AIS and categorize the stroke subtype into one of 4 mutually exclusive categories: definite arteriopathy, probable arteriopathy, cardioembolic, and idiopathic. While probable arteriopathy included cases of abnormal vascular imaging of uncertain etiology (arteriopathic vs. thromboembolic), the definite arteriopathy category was reserved for a high degree of diagnostic certainty. Given the probable arteriopathy group was likely very heterogeneous, this group was excluded from stroke subtype analyses.

**eMethods2: Differential Gene Expression Bioinformatics Pipeline**

To generate raw human transcript counts, raw fastq sequence files were run through a computational pipeline that incorporates quality filtering using paired-read iterative contig extension (PRICE)^23^ followed by human gene alignment using the Spliced Transcripts Alignment to a Reference (STAR) (v2.5.3a)^24^ as previously described.^25,26^ DGE analysis was performed using the EdgeR (version 4.4.2) package in R (version 4.4.2). Data preprocessing included removal of transcripts aligning to non-protein coding genes, removal of transcripts aligning to lowly expressed genes via filterByExp (retaining genes with ~10 raw counts in at least two samples), and trimmed mean of M (TMM) normalization. Normalized expression counts were fit to gene-wise generalized linear models based on the negative binomial distribution using a design matrix that incorporated sequencing batch, sex, and age strata as *a priori* suspected co-variates, to evaluate DGE between groups. A false discovery rate (FDR), generated by the Benjamini-Hochberg (BH) p-value correction for multiple hypothesis testing, of <0.05 and a log 2-fold change (log_2_FC) of greater than |1| was used to select the final list of differentially expressed genes.

**eFigure1: GO Pathway analysis of upregulated genes in all AIS versus stroke-free well controls**

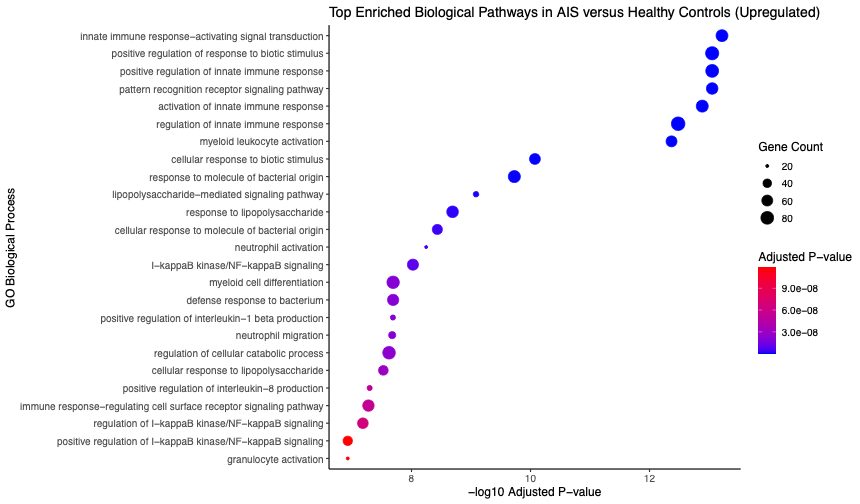

**eFigure2: GO Pathway analysis of downregulated genes in all AIS versus stroke-free well children**

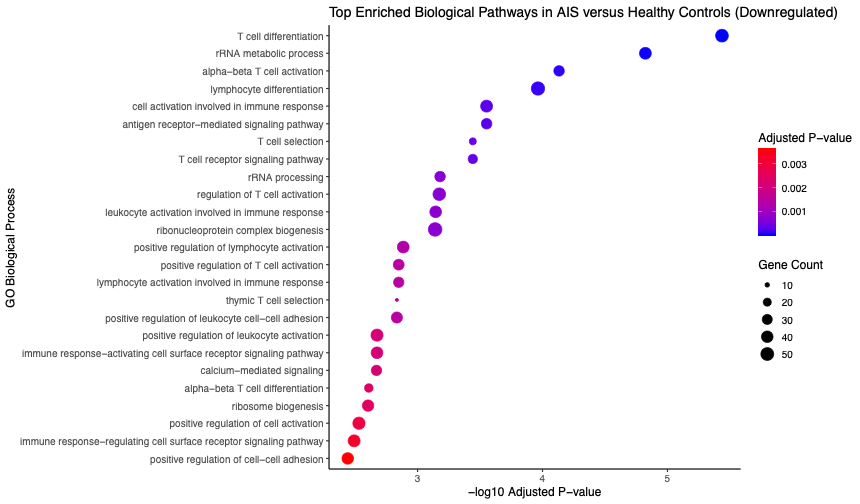

**eFigure3: GSEA of all genes in AIS versus stroke-free well children**

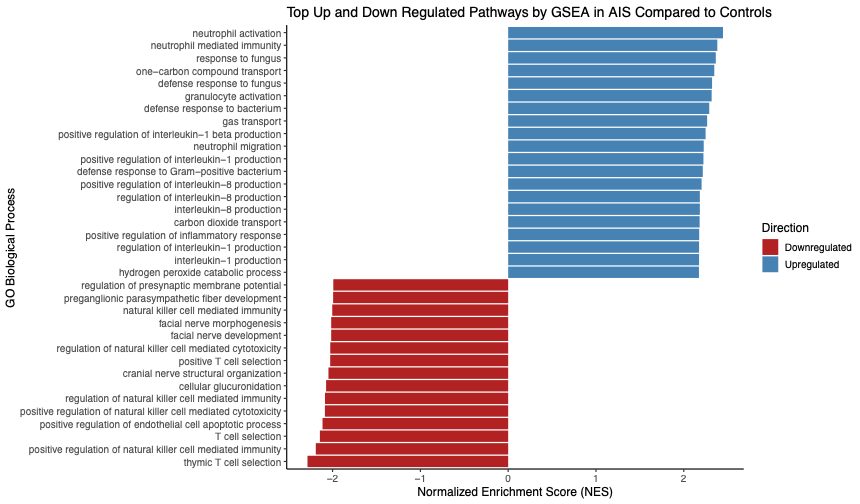

**eFigure4: GO Pathway of upregulated genes in Arteriopathic AIS versus stroke-free well children**

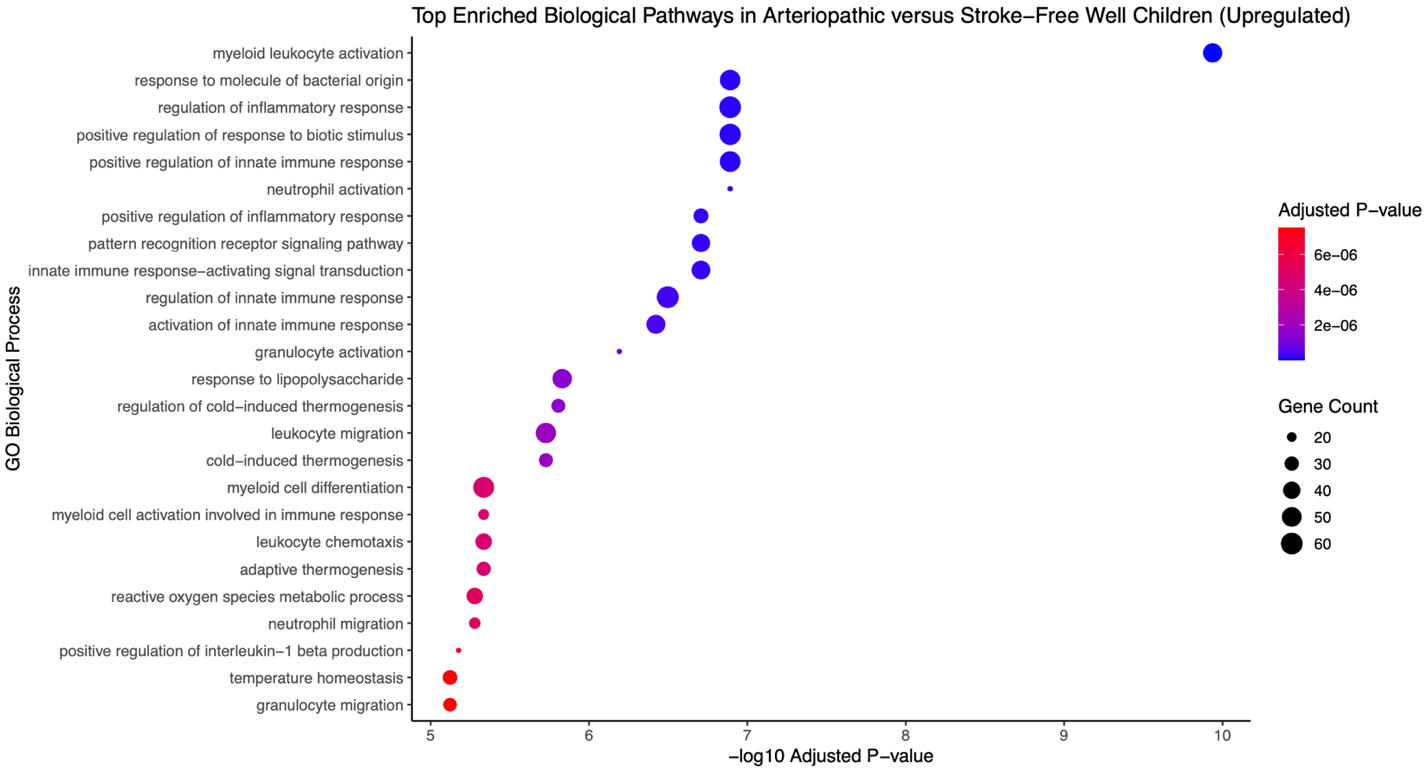

**eFigure5: GO Pathway of upregulated genes in Cardioembolic AIS versus stroke-free well children**

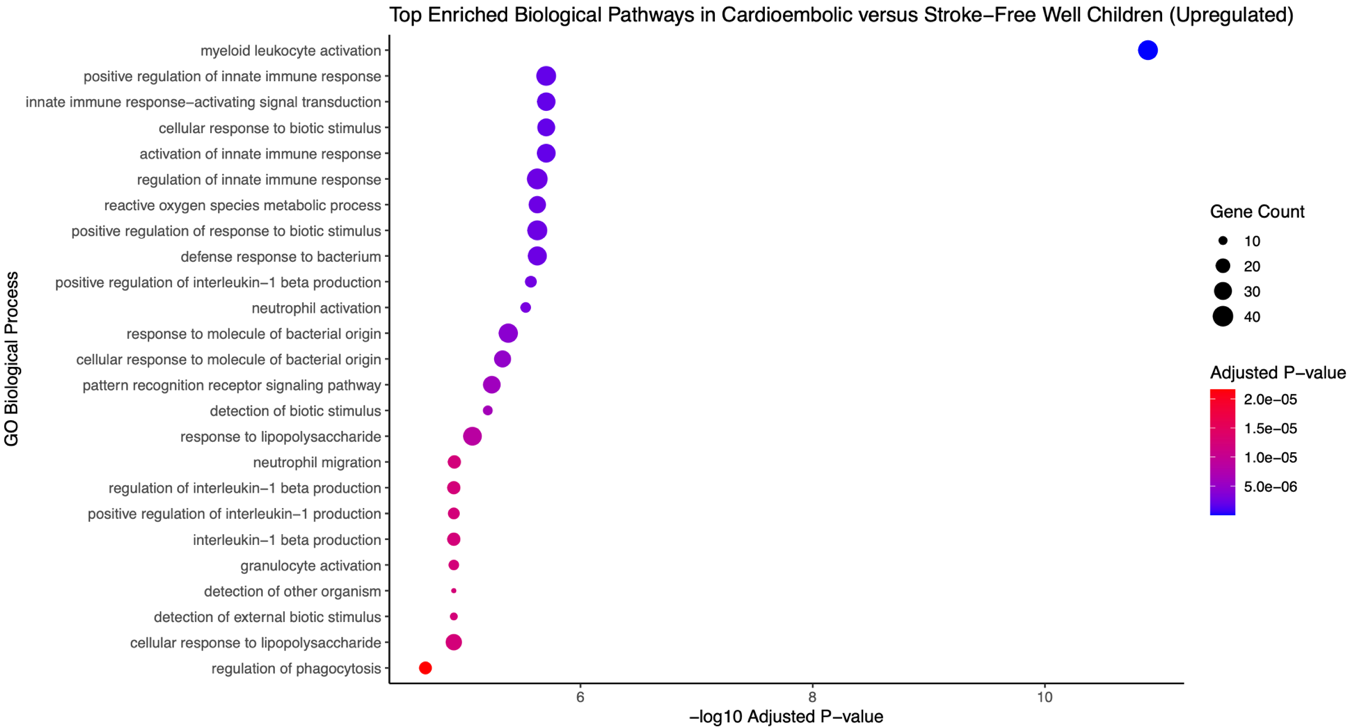

**eFigure6: Volcano plot of differentially expressed genes in arteriopathic vs. idiopathic AIS**

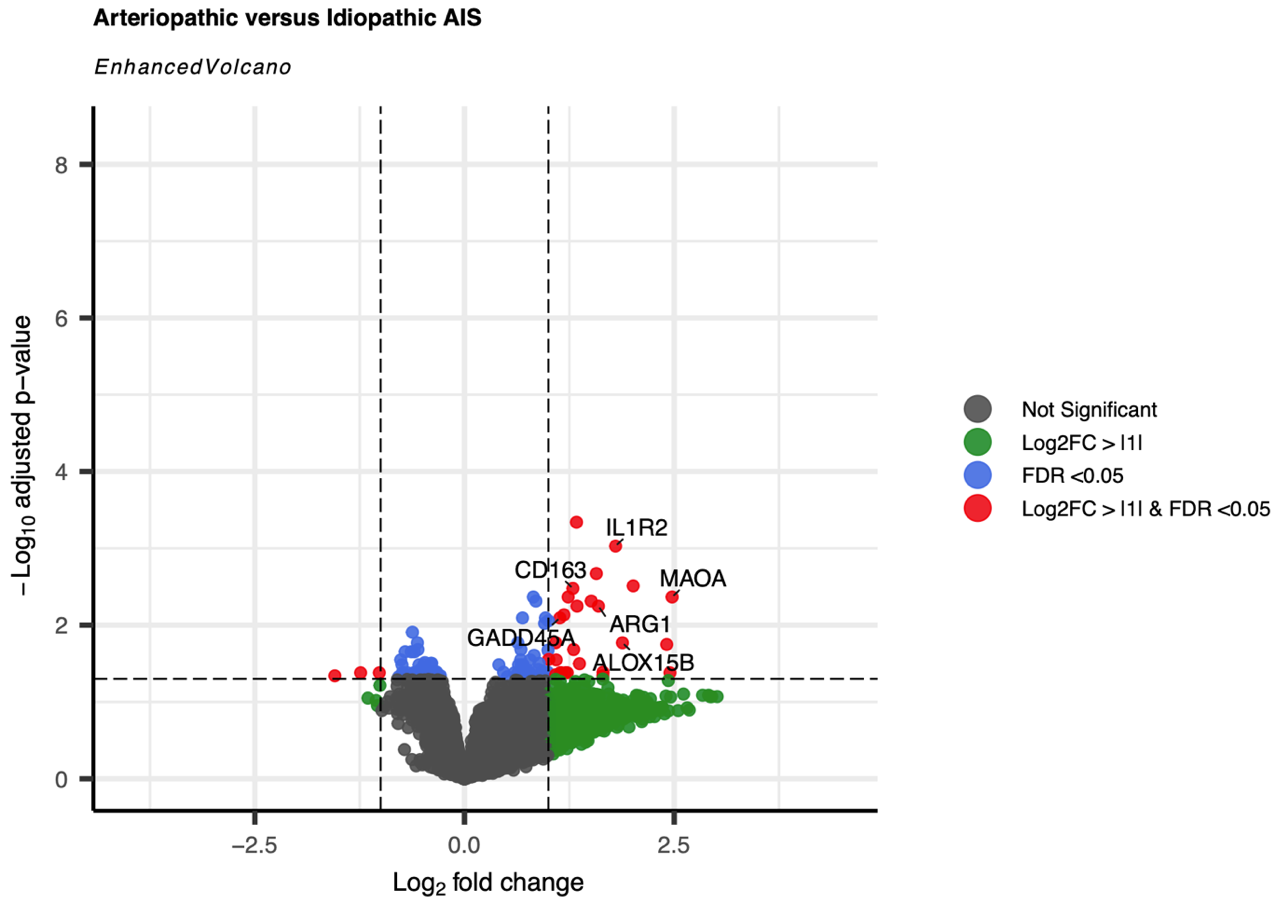

**eFigure7: Volcano plot of differentially expressed genes in cardioembolic vs. idiopathic AIS**

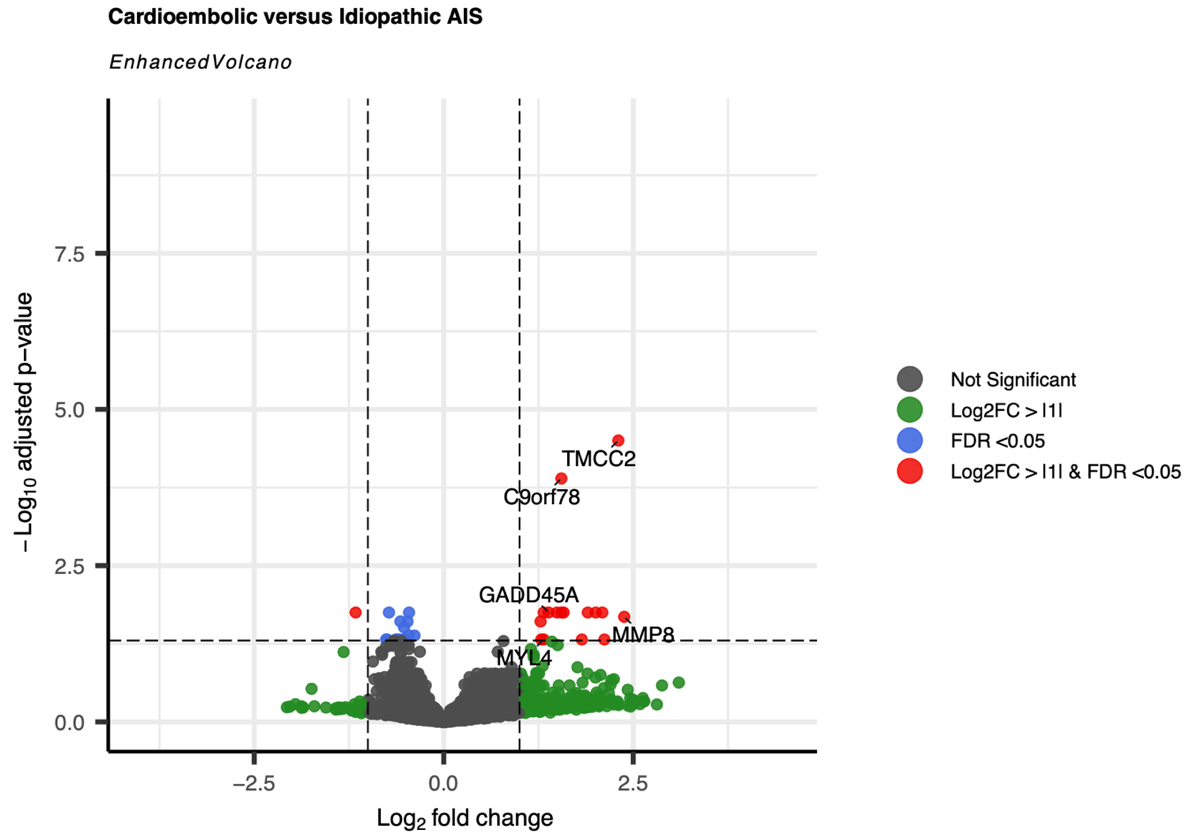

**eFigure8: Volcano plot of differentially expressed genes in arteriopathic vs. cardioembolic AIS**

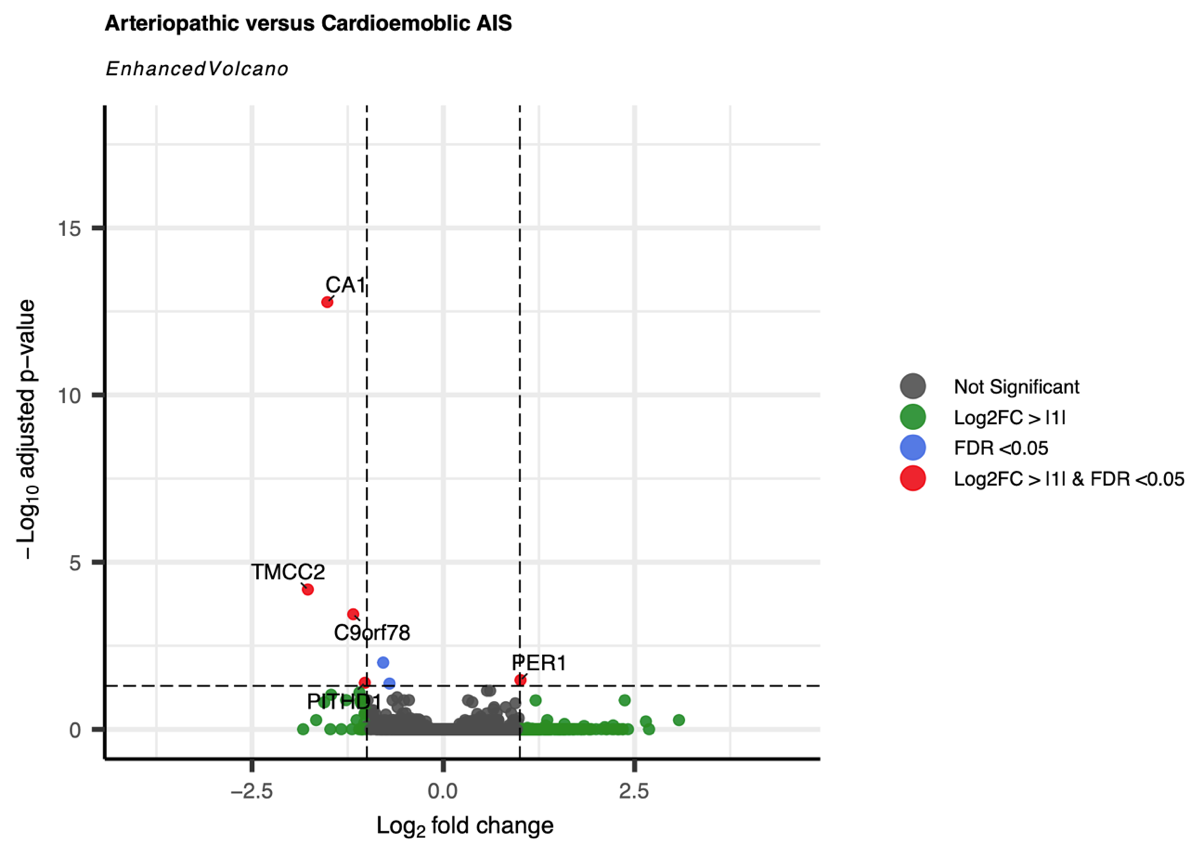
